# Defining the Burden of Disease of RSV in Europe: estimates of RSV-associated hospitalisations in children under 5 years of age. A systematic review and modelling study

**DOI:** 10.1101/2023.02.10.23285756

**Authors:** Marco Del Riccio, Peter Spreeuwenberg, Richard Osei-Yeboah, Caroline K. Johannesen, Liliana Vazquez Fernandez, Anne C. Teirlinck, Xin Wang, Terho Heikkinen, Mathieu Bangert, Saverio Caini, Harry Campbell, John Paget, the RESCEU investigators

## Abstract

**Background:** Respiratory syncytial virus (RSV) infections cause an estimated 22-50 million episodes of acute lower respiratory infections (ALRI) every year in children younger than 5 years. To date, no overall estimate of RSV-associated hospitalisations in children under 5 years has been published for the European Union (EU). Through statistical modelling, we estimated the RSV hospitalisation burden in children under 5 years of age in EU countries and Norway, by age group and country.

**Methods:** We collated national RSV-associated hospitalisation estimates calculated using linear regression models in children under 5 years via the RESCEU project for Denmark, England, Finland, Norway, the Netherlands and Scotland during 2006-2018. A systematic literature review was conducted to collect additional estimates. Using the multiple imputation and nearest neighbour matching extrapolation methods, we estimated RSV-associated hospitalisation rates in all EU countries.

**Results:** Additional estimates for Spain and France were found in the literature and added to the analysis. We estimated that an average of 245,244 (95%CI 224,688-265,799) hospital admissions with a respiratory infection per year were associated with RSV in children under the age of 5, with most cases occurring among children aged less than 1 year (75%). Infants aged less than 2 months represented the most affected group (71.6 per 1,000 children; 95%CI: 66.6-76.6). The hospitalisation rates varied widely across countries: for example, estimated rates in the 0-2 months age group ranged from 47.4 (37.5-57.3) per 1,000 in the Netherlands to 98.3 (88.5-108.1) per 1,000 in France.

**Conclusion:** To our knowledge, this is the first attempt to estimate the overall RSV hospitalisation burden in children under the age of 5 years in the EU. Our findings will help support decisions regarding prevention efforts, and they will also represent an important benchmark to understand changes in the RSV burden following the introduction of RSV immunisation programs in Europe.

## Introduction

It is globally estimated that respiratory syncytial virus (RSV) is associated with about 22% of all acute lower respiratory infections (ALRI) [1] and this results in approximately 101,400 (84,500 – 125,200) deaths per year in young children [2]. Several studies have been conducted to understand the burden of RSV-associated infections, hospitalisations, and deaths in children in Europe. For example, Reeves et al. explored routinely collected hospital data on RSV in children aged <5 years in 7 European countries and compared these to RSV-associated admission rates [3], while Demont et al. provided information on the clinical and economic burden of RSV-associated hospitalisation in children aged <5 years in France between 2010 and 2018 [4]. Despite these efforts, no estimates for RSV-associated hospitalisations are available for children in the EU as a whole.

RSV-associated hospitalisation estimates are important for public health purposes, as they can help allocate resources, and provide important insights and inputs for prevention measures and strategies. The establishment of a robust age-specific burden of disease estimates, which have often been limited due to a lack of routine testing for RSV [5], has also been underlined by the World Health Organization (WHO) [6].

In this paper, we present overall and country-specific estimates of RSV-associated respiratory hospitalisations (absolute numbers and rates) by age group in children aged less than 5 years in the 28 countries of the European Union (EU-28: includes the United Kingdom (UK) as it was part of EU when the data were collected) and Norway. We then used these estimates to calculate the proportion of RSV-associated hospitalisations among all-cause hospitalisations and respiratory hospitalisations in this age group, for each country. Estimates of the hospital burden of RSV are not available in many EU countries and our data will allow comparisons between countries and with other regions of the world, support efforts to communicate the RSV disease burden, and provide important data for decisions regarding future prevention and control measures linked to various immunisation programmes (such as vaccines and/or monoclonal antibodies).

## Methods

### Data sources

We searched for published and unpublished national estimates of RSV-associated hospitalisations (defined as any admission that contained at least one respiratory infection-specific ICD-10 code at any point during admission) in children under 5 years in EU countries that were calculated using regression models as input data for the statistical analysis.

### RSV-associated hospitalisation estimates from the RESCEU project

The data sources for Denmark, England, Finland, Norway, the Netherlands, and Scotland have been described in papers that were previously published by the REspiratory Syncytial virus Consortium in EUrope (RESCEU) [3]. Briefly, a retrospective study of overall respiratory hospital admissions (i.e., respiratory tract infections with or without an associated pathogen), RSV-related respiratory admissions, and other pathogen-respiratory admissions in children <5 years of age using routinely collected hospital admissions databases was conducted in these 6 countries of the EU/European Economic Area (EEA). Age-specific estimates of RSV-associated hospitalisations in children during 2006-2018 were calculated for these countries by using a linear regression approach [7], with estimates available for the following age groups: 0-2, 3-5, 6-11, 12-35, and 36-59 months.

### Literature review estimates to identify estimates in other countries

In order to increase the geographical representativeness of this work, we searched the scientific literature for additional data points in EU countries by adopting the same search strategy as a previously published systematic review that aimed to estimate the global incidence, hospital admission rate, and mortality due to RSV in young children based on national estimates [2]. The systematic review by Li et al. was broader in scope and inclusion criteria (i.e., they included studies reporting incidence and in-hospital and out-of-hospital mortality, not only hospitalisation rates), therefore, we considered that the included records needed to be further screened and assessed for eligibility in our review.

The new search was conducted according to the Preferred Reporting Items for Systematic Reviews and Meta-Analyses (PRISMA) statement [8]. The same search string as the systematic review conducted by Li and colleagues was used [2] (see Supplementary material); MEDLINE and EMBASE databases were searched from 1^st^ January 2019 to 30^th^ November 2021 for original articles. Papers published before 2019 that had been included by Li and colleagues were added to the reference list of relevant papers and further assessed for eligibility. No language restrictions were applied as long as an English abstract was available to decide on eligibility. To be included, a study had to report (i) national estimates of RSV-associated hospitalisations in children in EU countries calculated by using regression models (in order to have a homogeneous pool of estimates) (ii) details (i.e., ICD codes) on the diagnosis (i.e., bronchiolitis, lower respiratory infections, etc.), and (iii) analyse the same age bands used by Johannesen and colleagues [7].

After removing duplicates, titles and abstracts were independently screened by two researchers (MDR and ROY) and the articles that were not excluded were retrieved in full copy and independently read by two authors (MDR and ROY). The papers that had been included in the previous systematic review were further assessed for eligibility, and the reference list of all eligible papers was checked by means of backward citation chaining for further relevant references. Data extraction was organised using an internally piloted spreadsheet [2]. The estimates reported by the included articles were then extracted and used as input data for the statistical modelling, along with other information (data sources, details on the primary diagnosis, years in which the study was conducted, etc.). The quality assessment of the included studies was conducted by using a tool designed by Li et al. [2] (see Supplementary material). Based on the assessment of different questions (on study testing, subjects, case definition, sampling strategy, diagnostic tests, adjustment for health-care utilization) an overall score was calculated.

### Statistical Analysis

A two stage-modelling approach was used to estimate the respiratory hospitalisation burden of RSV in children under 5 years old. This method was adapted from prior work focused on influenza-associated mortality during the 2009 pandemic [9] and for seasonal influenza [10]. Since the period covered by the eligible studies was 2006-2018, the United Kingdom (UK) was included in the EU estimates.

In Stage 1, we identified annual age-specific estimates of RSV-associated hospitalisations from respiratory causes that were calculated using regression models. Data from the six RESCEU countries, plus those which were found in the literature and matched the inclusion criteria (see Results), were used as input data for Stage 2.

In Stage 2, we used the country estimates to extrapolate the hospitalisation burden and generate plausible values for all EU countries and Norway using two different modelling approaches, each involving two steps: (1) a data creation step using the matching approach or the multiple imputation approach, and (2) a data analysis step where a hierarchical linear random effects model is used to project the burden in all EU countries. For both approaches, the data creation step relied on 10 country-specific indicators representing health conditions at a demographic, geographic and population level (see Supplementary material) [9]. As two different sets of 10 indicators were used, the statistical modelling produced 4 sets of results (see Supplementary material), each related to the combination of one set of indicators and one modelling approach. An average of the four different models was calculated and this was used to calculate the absolute annual number and rates of RSV-associated hospitalisations (with uncertainty intervals) by country for the following age groups: 0-2, 3-5, 6-11, 12-35, and 36-59 months, which are consistent with the previously published age groups used by Johannesen and colleagues [7]. We also assessed the hospitalisations rate for EU28 in children aged 0-59 months and calculated the ratio of hospitalisations that occurred in each age group, considering our estimated total number of RSV-associated hospitalisations as denominator.

Finally, in order to compare RSV-associated hospitalisations to total respiratory and all-cause hospitalisations, the estimated absolute number of RSV-associated hospitalisations was used as the numerator to calculate the proportion of RSV-associated hospitalisations among respiratory and all-cause hospitalisations (data related to 2015) occurring in children under 5 years. The population denominator (roughly 25,900,000 children under 59 months of age in EU28 in 2015) and other demographic indicators used for the analysis were obtained from Eurostat [11]. Statistical analyses were conducted using Stata version 16 (Stata Corp, College Station, TX).

## Results

### Results of the literature review and Stage 1

The literature search in MEDLINE and EMBASE produced 1,372 unique entries, and an additional 33 articles were found by backward citation chaining or because they had been included by Li and colleagues in the previous systematic review [2] (Figure 1). Of these, 1,304 were excluded based on title and abstract and 101 were read in full text: 99 were excluded for not matching the inclusion criteria; the main reason for exclusion was not presenting national RSV-associated hospitalisation estimates calculated by using regression methods. Two studies, in particular, were excluded as they focused on different age groups [12–13] but they provided useful estimates as they covered a country (England) that was included in Stage 1 (estimates for England were already available) and were therefore comparable to the data reported by Johannesen et al [7]. Finally, two studies [14–15] reporting RSV-associated hospitalisation estimates for Spain from 1997 to 2011 and for France during 2010-2018 were included in the review and their estimates were used as input data for the statistical modelling (Stage 2).

**Figure 1:**
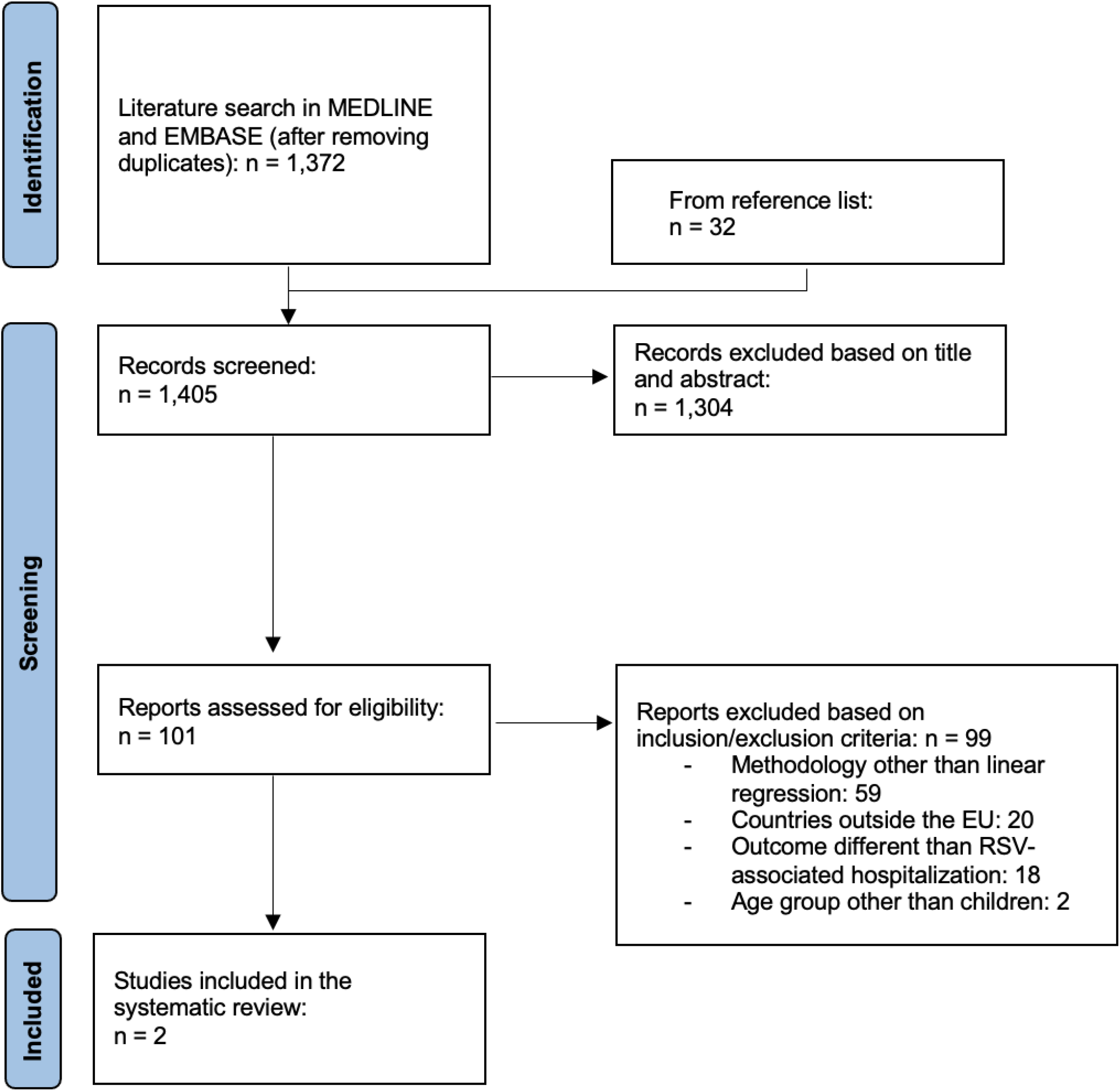
Systematic review flow diagram

**Figure 2:**
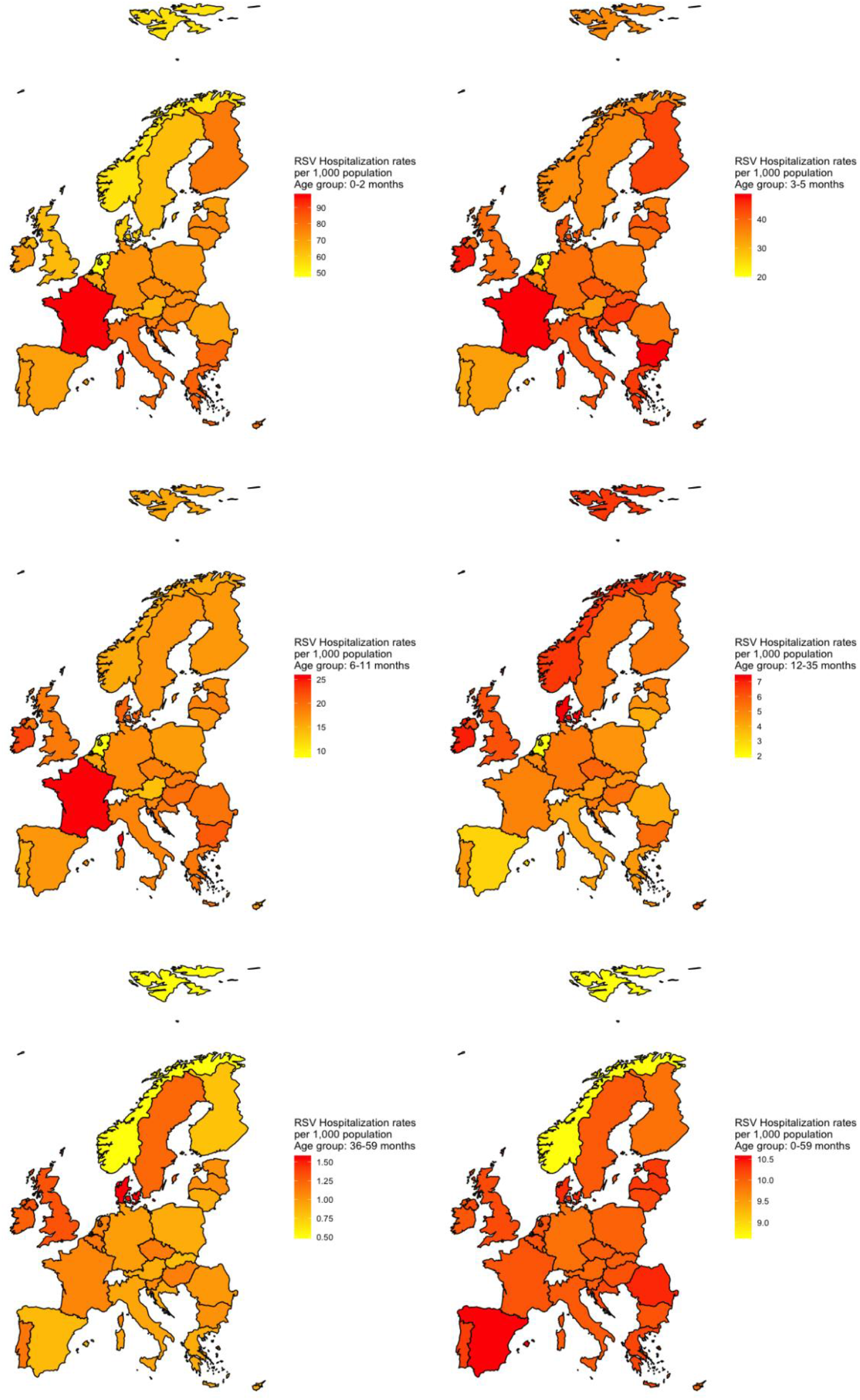
RSV Hospitalization rates per 1,000 population in different age groups

Both studies reported RSV-associated hospitalisation rates per 100,000 children in age groups that were slightly different to those used by Johannesen et al.; we therefore only used data related to the age groups that were consistent with the estimates produced by Johannesen et al. [7] (12-35 months and 36-59 months for Spain and 0-2 months, 3-5 months, and 6-11 months for France). Moreover, the estimates for France were not annual but based on the epidemic periods only (October to March), so we recalculated the estimates for the whole year by arbitrarily assuming little RSV activity during April-September (10% of the activity observed from October to March [16]).

In total, data from 8 countries were used as Stage 1 inputs [(Denmark, England, Finland, Norway, Netherlands, Scotland, France (age groups 0-2 months, 3-5 months, 6-11 months), and Spain (age groups 12-35 months, 36-59 months)] (Table 1).

**Table 1.** Description of the data sources that provided Stage 1 estimates

| Author, year | Country | Period of observation | Age groups | Age groups whose estimates were used as inputs | Outcome coding |
| --- | --- | --- | --- | --- | --- |
| Johannesen, 2022 [7] | Denmark | 2010-2017 | 0-2m, 3-5m, 6-11m, 12-35m, 36-59m | 0-2m, 3-5m, 6-11m, 12-35m, 36-59m | ICD-10; J00, J02-06 acute upper respiratory tract infection (URTI); J09-18 Pneumonia & influenza; J20-21, J40 Bronchiolitis and bronchitis; J22 Unspecified LRTI |
| Johannesen, 2022 [7] | England | 2007-2017 | 0-2m, 3-5m, 6-11m, 12-35m, 36-59m | 0-2m, 3-5m, 6-11m, 12-35m, 36-59m | ICD-10; J00, J02-06 acute upper respiratory tract infection (URTI); J09-18 Pneumonia & influenza; J20-21, J40 Bronchiolitis and bronchitis; J22 Unspecified LRTI |
| Johannesen, 2022 [7] | Finland | 2006-2016 | 0-2m, 3-5m, 6-11m, 12-35m, 36-59m | 0-2m, 3-5m, 6-11m, 12-35m, 36-59m | ICD-10; J00, J02-06 acute upper respiratory tract infection (URTI); J09-18 Pneumonia & influenza; J20-21, J40 Bronchiolitis and bronchitis; J22 Unspecified LRTI |
| Johannesen, 2022 [7] | Netherlands | 2013-2017 | 0-2m, 3-5m, 6-11m, 12-35m, 36-59m | 0-2m, 3-5m, 6-11m, 12-35m, 36-59m | ICD-10; J00, J02-06 acute upper respiratory tract infection (URTI); J09-18 Pneumonia & influenza; J20-21, J40 Bronchiolitis and bronchitis; J22 Unspecified LRTI |
| Johannesen, 2022 [7] | Norway | 2008-2017 | 0-2m, 3-5m, 6-11m, 12-35m, 36-59m | 0-2m, 3-5m, 6-11m, 12-35m, 36-59m | ICD-10; J00, J02-06 acute upper respiratory tract infection (URTI); J09-18 Pneumonia & influenza; J20-21, J40 Bronchiolitis and bronchitis; J22 Unspecified LRTI |
| Johannesen, 2022 [7] | Scotland | 2010-2016 | 0-2m, 3-5m, 6-11m, 12-35m, 36-59m | 0-2m, 3-5m, 6-11m, 12-35m, 36-59m | ICD-10; J00, J02-06 acute upper respiratory tract infection (URTI); J09-18 Pneumonia & influenza; J20-21, J40 Bronchiolitis and bronchitis; J22 Unspecified LRTI |
| Demont, 2020 [15] | France | 2010-2018 | 0-2m, 3-5m, 6-11m, 12-23m, 24-59m | 0-2m, 3-5m, 6-11m | ICD-10; J121, J205, J210, J219 |
| Gil-Prieto, 2015 [14] | Spain | 1997-2011 | 0y, 1y, 2y, 3y, 4y, <5y, <2y | 2y, 3y, 4y | ICD-9-CM; 466, acute bronchitis and bronchiolitis; 480.1, pneumonia due to RSV; 079.6, RSV infection |

### Stage 2 estimates

We included Stage 1 estimates from 8 countries in our analysis, roughly representing 40% of the population of the EU and Norway [17]. The results produced by the four models were consistent across the age groups, the highest rates being calculated with the multiple imputation approach and the lowest rates being calculated with the nearest neighbour matching approach (5% variation between the highest and lowest estimates of RSV hospitalisation rates in children aged 0-2 months) (see Supplementary material). Here, we present the results for the average of the four models.

We estimated the average number and average annual rate (per 1,000 population) of RSV-associated hospitalisations by age group in different countries (Table 2 and Table 3). We estimated that an average of 245,244 (95%CI 224,688-265,799) hospital admissions with respiratory infection were associated with RSV in the 28 EU countries per year in children under the age of 5, with most cases occurring among children aged less than 1 year (74.9%) and those aged 1-2 years (20.7%) (Table 4). Infants aged less than 2 months represented the most affected group (71.6 per 1,000 population; 95%CI: 66.6-76.6), with the rates declining as the children got older: 38.9 per 1,000 in children aged 3-5 months, 17.6 (6-11 months), 5.0 (12-35 months) and 1.0 (36-59 months). Overall, we estimated that an average of 10 children per 1,000 living in the European Union are hospitalized due to RSV annually (average rates in children 0-59 months: 10.06, 9.90-10.21 per 1,000 population).

**Table 2:** Average RSV-associated hospitalisation rates per 1,000 population per age group per year

| Country | 0-2 months (95%CI)<sup>a</sup> | 3-5 months (95%CI)<sup>a</sup> | 6-11 months (95%CI)<sup>a</sup> | 12-35 months (95%CI)<sup>b</sup> | 36-59 months (95%CI)<sup>b</sup> |
| --- | --- | --- | --- | --- | --- |
| EU28<sup>c</sup> | 71.6 (66.6-76.6) | 38.9 (36-41.9) | 17.6 (16.1-19.1) | 5 (4.4-5.5) | 1 (0.9-1.1) |
| Austria | 65 (55.2-74.8) | 33.1 (27.7-38.5) | 13.5 (10.7-16.4) | 4.5 (3.5-5.5) | 0.9 (0.7-1.1) |
| Belgium | 68.6 (58.8-78.4) | 36.3 (30.9-41.7) | 15.9 (13.1-18.7) | 4.8 (3.8-5.9) | 1.2 (0.9-1.4) |
| Bulgaria | 81.8 (72-91.6) | 48.9 (43.5-54.3) | 21.2 (18.4-24) | 5.5 (4.5-6.6) | 1 (0.8-1.2) |
| Croatia | 79.4 (69.7-89.2) | 42.6 (37.2-48) | 19 (16.2-21.8) | 4.8 (3.8-5.8) | 0.9 (0.7-1.1) |
| Cyprus | 78.4 (68.6-88.2) | 41.3 (35.9-46.7) | 16.7 (13.9-19.5) | 5.5 (4.5-6.5) | 0.9 (0.7-1.1) |
| Czech Republic | 73.9 (64.1-83.7) | 41 (35.6-46.4) | 18.4 (15.6-21.2) | 5.7 (4.6-6.7) | 1.1 (0.9-1.3) |
| Denmark | 59.2 (49.3-69) | 40.7 (35.3-46.2) | 20.4 (17.5-23.2) | 7.5 (6.4-8.5) | 1.6 (1.4-1.8) |
| Estonia | 69.8 (60-79.7) | 37.3 (31.8-42.7) | 17.2 (14.4-20.1) | 5.1 (4-6.1) | 1 (0.8-1.2) |
| Finland | 77.8 (67.9-87.7) | 43.2 (37.7-48.6) | 16.6 (13.8-19.4) | 5.2 (4.2-6.3) | 0.8 (0.6-1) |
| France | 98.3 (88.5-108.1) | 48.8 (43.4-54.2) | 26 (23.2-28.8) | 5 (4-6.1) | 1.1 (0.9-1.3) |
| Germany | 72.5 (62.7-82.2) | 38.6 (33.2-44) | 17.5 (14.6-20.3) | 5.2 (4.2-6.3) | 1 (0.8-1.2) |
| Greece | 82.6 (72.8-92.4) | 44.3 (38.9-49.7) | 19.1 (16.3-21.9) | 4.5 (3.4-5.5) | 0.9 (0.7-1.1) |
| Hungary | 75.3 (65.5-85.1) | 44.7 (39.3-50.1) | 19.8 (16.9-22.6) | 5.4 (4.4-6.4) | 1.1 (0.9-1.3) |
| Ireland | 70.1 (60.3-79.9) | 47 (41.6-52.4) | 22.6 (19.8-25.4) | 7.1 (6-8.1) | 1.3 (1-1.5) |
| Italy | 80.9 (71.1-90.7) | 41.7 (36.3-47.1) | 18.1 (15.3-20.9) | 4.3 (3.2-5.3) | 0.9 (0.7-1.1) |
| Latvia | 75 (65.2-84.8) | 41.3 (35.9-46.7) | 18.4 (15.6-21.2) | 4.6 (3.6-5.7) | 1.1 (0.8-1.3) |
| Lithuania | 73.6 (63.8-83.4) | 38.5 (33.1-43.9) | 17 (14.2-19.8) | 4 (3-5) | 0.9 (0.7-1.1) |
| Luxembourg | 63.5 (53.7-73.3) | 32.6 (27.2-38) | 13.1 (10.3-15.9) | 5 (4-6.1) | 0.9 (0.7-1.1) |
| Malta | 64.8 (54.8-74.7) | 35.2 (29.7-40.7) | 17.9 (15-20.7) | 5.4 (4.4-6.5) | 1.3 (1.1-1.5) |
| Netherlands | 47.4 (37.6-57.3) | 19.9 (14.5-25.4) | 8.5 (5.7-11.3) | 1.9 (0.8-2.9) | 1.1 (0.9-1.3) |
| Norway | 54.6 (44.5-64.7) | 34.8 (29.3-40.3) | 15.4 (12.5-18.2) | 6.6 (5.6-7.7) | 0.5 (0.3-0.7) |
| Poland | 71.4 (61.6-81.2) | 36.5 (31.1-41.9) | 16.2 (13.4-19.1) | 4.6 (3.5-5.6) | 0.9 (0.7-1.1) |
| Portugal | 70.2 (60.4-80) | 31.7 (26.3-37.1) | 15.2 (12.4-18) | 4.5 (3.4-5.5) | 1.2 (0.9-1.4) |
| Romania | 68.8 (59-78.6) | 37.7 (32.3-43.1) | 19.4 (16.6-22.2) | 4.1 (3.1-5.2) | 1 (0.8-1.2) |
| Slovakia | 74.6 (64.8-84.3) | 42 (36.6-47.4) | 18.8 (16-21.6) | 4.7 (3.7-5.8) | 0.8 (0.6-1) |
| Slovenia | 75.4 (65.6-85.2) | 42.6 (37.2-48) | 17.6 (14.8-20.4) | 5.2 (4.2-6.2) | 1.1 (0.9-1.3) |
| Spain | 69.4 (59.6-79.2) | 32.6 (27.2-38) | 16.8 (14-19.6) | 3 (2-4.1) | 0.8 (0.6-1) |
| Sweden | 63 (53.1-72.8) | 35.5 (30.1-41) | 16.9 (14.1-19.7) | 5.3 (4.3-6.3) | 1.2 (1-1.4) |
| United Kingdom | 63.4 (47.8-79.1) | 38.9 (29.7-48.1) | 18.9 (14.2-23.6) | 6.1 (4.4-7.9) | 1.3 (1-1.6) |

<sup>a</sup> RSV-associated hospitalisation rates in these three age groups for the 29 countries are estimated by also including data from France reported by Demont and
colleagues [15]
<sup>b</sup> RSV-associated hospitalisation rates in these two age groups for the 29 countries are estimated by also including data from Spain reported by Gil-Prieto and colleagues
<sup>c</sup> Includes the UK and excludes Norway

**Table 3:** Average RSV-associated hospitalisations per age group per year

| Country | 0-2 months (95%CI)<sup>a</sup> | 3-5 months (95%CI)<sup>a</sup> | 6-11 months (95%CI)<sup>a</sup> | 12-35 months (95%CI)<sup>b</sup> | 36-59 months (95%CI)<sup>b</sup> |
| --- | --- | --- | --- | --- | --- |
| EU28<sup>c</sup> | 90,200 (83,923-96,476) | 49,052 (45,328-52,776) | 44,369 (40,529-48,208) | 50,852 (45,249-56,456) | 10,771 (9,659-11,883) |
| Austria | 1,308 (1,111-1,505) | 667 (558-775) | 545 (432-658) | 732 (563-902) | 147 (112-182) |
| Belgium | 2,141 (1,836-2,446) | 1,133 (965-1,302) | 992 (816-1,167) | 1,235 (973-1,497) | 306 (250-362) |
| Bulgaria | 1,374 (1,210-1,539) | 822 (732-913) | 714 (620-808) | 733 (596-869) | 141 (112-171) |
| Croatia | 783 (687-880) | 420 (366-473) | 375 (320-431) | 391 (306-477) | 77 (59-95) |
| Cyprus | 181 (159-204) | 95 (83-108) | 77 (64-90) | 106 (86-126) | 18 (13-22) |
| Czech Republic | 2,031 (1,762-2,300) | 1,128 (979-1,276) | 1,012 (858-1,167) | 1,237 (1,012-1,462) | 258 (209-308) |
| Denmark | 846 (704-986) | 582 (504-660) | 582 (500-662) | 864 (744-985) | 199 (172-225) |
| Estonia | 238 (205-271) | 127 (108-146) | 118 (98-137) | 141 (112-170) | 32 (25-38) |
| Finland | 1,122 (980-1,264) | 622 (544-701) | 479 (398-561) | 625 (501-750) | 97 (71-124) |
| France | 18,145 (16,336-19,952) | 9,018 (8,021-10,015) | 9,587 (8,548-10,626) | 7,573 (5,998-9,148) | 1,704 (1,368-2,040) |
| Germany | 12,977 (11,223-14,731) | 6,906 (5,939-7,874) | 6,252 (5,244-7,260) | 7,250 (5,802-8,699) | 1,334 (1,039-1,629) |
| Greece | 1,895 (1,670-2,119) | 1,015 (891-1,139) | 875 (746-1,003) | 862 (661-1,063) | 193 (147-239) |
| Hungary | 1,748 (1,521-1,975) | 1,038 (913-1,163) | 917 (787-1,048) | 976 (788-1,164) | 199 (161-237) |
| Ireland | 1,139 (980-1,298) | 764 (676-851) | 735 (643-826) | 950 (810-1,091) | 176 (146-206) |
| Italy | 10,111 (8,888-11,334) | 5,213 (4,538-5,888) | 4,534 (3,832-5,236) | 4,475 (3,387-5,563) | 1,021 (787-1,256) |
| Latvia | 407 (353-459) | 224 (195-253) | 200 (169-230) | 190 (148-232) | 40 (32-48) |
| Lithuania | 559 (485-633) | 292 (251-333) | 258 (215-300) | 241 (179-304) | 54 (42-67) |
| Luxembourg | 96 (82-111) | 49 (41-58) | 40 (31-49) | 64 (51-77) | 12 (9-15) |
| Malta | 69 (59-80) | 38 (32-44) | 38 (32-44) | 47 (38-57) | 11 (9-13) |
| Netherlands | 2,071 (1,641-2,502) | 870 (633-1,108) | 741 (494-988) | 651 (292-1,011) | 398 (319-475) |
| Norway | 811 (661-961) | 517 (435-599) | 456 (370-542) | 812 (682-941) | 61 (34-89) |
| Poland | 6,542 (5,646-7,439) | 3,346 (2,852-3,842) | 2,979 (2,464-3,494) | 3,459 (2,669-4,249) | 730 (557-903) |
| Portugal | 1,444 (1,243-1,645) | 651 (540-762) | 627 (511-742) | 769 (590-949) | 226 (185-268) |
| Romania | 3,300 (2,830-3,769) | 1,807 (1,548-2,066) | 1,860 (1,590-2,129) | 1,540 (1,150-1,929) | 389 (305-472) |
| Slovakia | 1,035 (900-1,170) | 583 (509-658) | 523 (445-600) | 531 (414-648) | 98 (72-123) |
| Slovenia | 399 (347-451) | 225 (197-254) | 186 (156-216) | 225 (180-270) | 48 (38-57) |
| Spain | 7,399 (6,356-8,442) | 3,473 (2,897-4,048) | 3,574 (2,975-4,172) | 2,670 (1,762-3,578) | 788 (585-992) |
| Sweden | 1,824 (1,538-2,110) | 1,030 (872-1,187) | 980 (816-1,144) | 1,229 (987-1,471) | 288 (237-339) |
| United Kingdom | 12,333 (9,291-15,375) | 7,565 (5,778-9,351) | 7,352 (5,515-9,188) | 9,890 (7,128-12,652) | 2,156 (1,614-2,698) |

<sup>a</sup> RSV-associated hospitalisation in these three age groups are estimated by also including data from France reported by Demont and colleagues [15]
<sup>b</sup> RSV-associated hospitalisation in these two age groups are estimated by also including data from Spain reported by Gil-Prieto and colleagues [14]
<sup>c</sup> Includes the UK and excludes Norway

**Table 4:** Ratio of RSV-associated hospitalisation occurring in children aged less than 1 year, from 1 to 2 years and from 3 to 4 years (100% is represented by all RSV-associated hospitalisation occurring in children under 5 years).

| Country | 0-2 months<sup>a</sup> - % | 3-5 months<sup>a</sup> - % | 6-11 months<sup>a</sup> - % | 0-11 months<sup>a,b</sup> - % | 12-35 months<sup>c</sup> - % | 36-59 months<sup>c</sup> - % |
| --- | --- | --- | --- | --- | --- | --- |
| EU28<sup>d</sup> | 36.8% | 20.0% | 18.1% | 74.9 | 20.7 | 4.4 |
| Austria | 38.5% | 19.6% | 16.0% | 74.1 | 21.6 | 4.3 |
| Belgium | 36.9% | 19.5% | 17.1% | 73.4 | 21.3 | 5.3 |
| Bulgaria | 36.3% | 21.7% | 18.9% | 76.9 | 19.4 | 3.7 |
| Croatia | 38.3% | 20.5% | 18.3% | 77.1 | 19.1 | 3.8 |
| Cyprus | 37.9% | 19.9% | 16.1% | 74.0 | 22.2 | 3.8 |
| Czech Republic | 35.8% | 19.9% | 17.9% | 73.6 | 21.8 | 4.6 |
| Denmark | 27.5% | 18.9% | 18.9% | 65.4 | 28.1 | 6.5 |
| Estonia | 36.3% | 19.4% | 18.0% | 73.6 | 21.5 | 4.9 |
| Finland | 38.1% | 21.1% | 16.3% | 75.5 | 21.2 | 3.3 |
| France | 39.4% | 19.6% | 20.8% | 79.8 | 16.5 | 3.7 |
| Germany | 37.4% | 19.9% | 18.0% | 75.3 | 20.9 | 3.8 |
| Greece | 39.2% | 21.0% | 18.1% | 78.2 | 17.8 | 4.0 |
| Hungary | 35.8% | 21.3% | 18.8% | 75.9 | 20.0 | 4.1 |
| Ireland | 30.3% | 20.3% | 19.5% | 70.1 | 25.2 | 4.7 |
| Italy | 39.9% | 20.6% | 17.9% | 78.3 | 17.7 | 4.0 |
| Latvia | 38.4% | 21.1% | 18.9% | 78.3 | 17.9 | 3.8 |
| Lithuania | 39.8% | 20.8% | 18.4% | 79.0 | 17.2 | 3.8 |
| Luxembourg | 36.8% | 18.8% | 15.3% | 70.9 | 24.5 | 4.6 |
| Malta | 34.0% | 18.7% | 18.7% | 71.4 | 23.2 | 5.4 |
| Netherlands | 43.8% | 18.4% | 15.7% | 77.8 | 13.8 | 8.4 |
| Norway | 30.5% | 19.5% | 17.2% | 67.1 | 30.6 | 2.3 |
| Poland | 38.4% | 19.6% | 17.5% | 75.4 | 20.3 | 4.3 |
| Portugal | 38.8% | 17.5% | 16.9% | 73.2 | 20.7 | 6.1 |
| Romania | 37.1% | 20.3% | 20.9% | 78.3 | 17.3 | 4.4 |
| Slovakia | 37.4% | 21.0% | 18.9% | 77.3 | 19.2 | 3.5 |
| Slovenia | 36.8% | 20.8% | 17.2% | 74.8 | 20.8 | 4.4 |
| Spain | 41.3% | 19.4% | 20.0% | 80.7 | 14.9 | 4.4 |
| Sweden | 34.1% | 19.2% | 18.3% | 71.6 | 23.0 | 5.4 |
| United Kingdom | 31.4% | 19.3% | 18.7% | 69.3 | 25.2 | 5.5 |

<sup>a</sup> RSV-associated hospitalisations in this age group are estimated by also including data from France reported by Demont and colleagues [15]
<sup>b</sup> Calculated as a total of the previous three age groups (0-2 months, 3-5 months, 6-11 months)
<sup>c</sup> RSV-associated hospitalisations in these two age groups are estimated by also including data from Spain reported by Gil-Prieto and colleagues [14]
<sup>d</sup> Includes the UK and excludes Norway

We also estimated country-specific RSV-associated respiratory hospitalisations for each of the 29 countries and rates per 1,000 children (Table 2 and Table 3). The countries which had the highest absolute number of estimated hospitalisations were France (46,027 hospitalisations per year in children under 5 years), the UK (39,296 hospitalisations), and Germany (34,719 hospitalisations).

The hospitalisation rates varied widely across the EU: in the first age group (0-2 months) they ranged from 47.4 (95%CI: 37.6-57.3) per 1,000 population in the Netherlands to 98.3 (88.5-108.1) in France. The Netherlands presented the lowest rates in almost all the other age groups: 19.9 (14.5-25.4) per 1,000 population in children aged 3-5 months, 8.5 (5.7-11.3) in children aged 6-11 months, 1.9 (0.8-2.9) in children aged 12-35 months. The lowest rates for the age group 36-59 months were estimated for Norway (0.5, 95%CI 0.3-0.7). In the age group 0-5 years (0-59 months), the rates ranged from 8.61 (8.31-8.92) in Norway to 10.58 (10.30-18.86) in Spain.

Most RSV-associated hospitalisations occurred in children aged less than 1 year (74.9% averaged, ranging from 65.4% in Denmark to 80.7% in Spain (Table 4)). The youngest group (0-2 months) was the most affected, with percentages ranging from 27.5% in Denmark to 43.8% in the Netherlands. RSV-associated hospitalisations were less likely in children aged from 3 to 4 years (36-59 months), with the percentage ranging from 3.3% in Finland to 8.4% in the Netherlands.

### Comparison with total pediatric hospitalisations and respiratory pediatric hospitalisations

We compared the country estimates to total national paediatric hospitalisations and respiratory paediatric hospitalisations in the EU and Norway and found that RSV-associated hospitalisations represented from 1.8% (95%CI: 1.5-2.1; Lithuania) to 9.9 (95%CI: 8.4-11.5; Finland) of total hospitalisations in children younger than 5 years (Table 5) [11]. This percentage was higher for paediatric respiratory hospitalisations, ranging from 6.8% in Lithuania to 51.6% in Sweden, and these percentages are likely to be much higher during the winter, especially during the weeks when RSV circulates.

**Table 5:** Hospitalizations in children under 5 years (all causes and respiratory causes) in EU28 countries and Norway, and % of all hospitalisations and all respiratory hospitalisations that were due to RSV.

| Country | Hospitalizations in children under 5 years, all causes<sup>a</sup> | Hospitalizations in children under 5 years, respiratory causes<sup>a</sup> | RSV-associated hospitalisations in children under 5 years<sup>b</sup> | % of all hospitalisations in children under 5 years due to RSV | % of all respiratory hospitalisations in children under 5 years due to RSV |
| --- | --- | --- | --- | --- | --- |
| Austria | na | 16,305 | 3,399 (2,776-4,022) |  | 20.8 (17.0-24.7) |
| Belgium | na | na | 5,807 (4,840-6,774) |  |  |
| Bulgaria | na | na | 3,784 (3,270-4,300) |  |  |
| Croatia | 62,972 | 8,060 | 2,046 (1,738-2,356) | 3.2 (2.8-3.7) | 25.4 (21.6-29.2) |
| Cyprus | 6,839 | 1,568 | 477 (405-550) | 7.0 (5.9-8.0) | 30.4 (25.8-35.1) |
| Czech Republic | 196,900 | 26,813 | 5,666 (4,820-6,513) | 2.9 (2.4-3.3) | 21.1 (18.0-24.3) |
| Denmark | 47,695 | 7,672 | 3,073 (2,624-3,518) | 6.4 (5.5-7.4) | 40.1 (34.2-45.9) |
| Estonia | na | na | 656 (548-762) |  |  |
| Finland | 29,637 | 6,854 | 2,945 (2,494-3,400) | 9.9 (8.4-11.5) | 43.0 (36.4-49.6) |
| France | 1,229,788 | 127,538 | 46,027 (40,271-51,781) | 3.7 (3.3-4.2) | 36.1 (31.6-40.6) |
| Germany | 1,253,873 | 162,515 | 34,719 (29,247-40,193) | 2.8 (2.3-3.2) | 21.4 (18.0-24.7) |
| Greece | na | na | 4,840 (4,115-5,563) |  |  |
| Hungary | 175,698 | 29,555 | 4,878 (4,170-5,587) | 2.8 (2.4-3.2) | 16.5 (14.1-18.9) |
| Ireland | 53,504 | 11,637 | 3,764 (3,255-4,272) | 7.0 (6.1-8.0) | 32.3 (28.0-36.7) |
| Italy | 731,993 | 62,922 | 25,354 (21,432-29,277) | 3.5 (2.9-4.0) | 40.3 (34.1-46.5) |
| Latvia | na | 9,608 | 1,061 (897-1,222) |  | 11.0 (9.3-12.7) |
| Lithuania | 78,166 | 20,663 | 1,404 (1,172-1,637) | 1.8 (1.5-2.1) | 6.8 (5.7-7.9) |
| Luxembourg | na | 777 | 261 (214-310) |  | 33.6 (27.5-39.9) |
| Malta | 7,030 | 642 | 203 (170-238) | 2.9 (2.4-3.4) | 31.6 (26.5-37.1) |
| Netherlands | na | 18,201 | 4,731 (3,379-6,084) |  | 26.0 (18.6-33.4) |
| Norway | 84,850 | 7,251 | 2,657 (2,182-3,132) | 3.1 (2.6-3.7) | 36.6 (30.1-43.2) |
| Poland | 664,693 | 120,409 | 17,056 (14,188-19,927) | 2.6 (2.1-3.0) | 14.2 (11.8-16.5) |
| Portugal | 98,167 | 8,069 | 3,717 (3,069-4,366) | 3.8 (3.1-4.4) | 46.1 (38.0-54.1) |
| Romania | 424,339 | 123,377 | 8,896 (7,423-10,365) | 2.1 (1.7-2.4) | 7.2 (6.0-8.4) |
| Slovakia | 113,651 | 19,071 | 2,770 (2,340-3,199) | 2.4 (2.1-2.8) | 14.5 (12.3-16.8) |
| Slovenia | 46,511 | 7,346 | 1,083 (918-1,248) | 2.3 (2.0-2.7) | 14.7 (12.5-17.0) |
| Spain | na | 58,598 | 17,904 (14,575-21,232) |  | 30.6 (24.9-36.2) |
| Sweden | na | 10,362 | 5,351 (4,450-6,251) |  | 51.6 (42.9-60.3) |
| United Kingdom | 979,392 | 116,819 | 39,296 (29,326-49,264) | 4.0 (3.0-5.0) | 33.6 (25.1-42.2) |

na: not available
<sup>a</sup> Data are related to 2015 [11]
<sup>b</sup> Estimates calculated in the present manuscript.

## Discussion

Understanding the burden of disease caused by RSV, and specifically the incidence of hospitalisations and deaths, will help understand the impact of RSV prevention programs (new monoclonal antibodies and vaccines [18–21]). Our study estimated that an average of roughly 250,000 respiratory hospitalisations in children younger than 5 years were associated with RSV each year in the 28 EU countries included in the analysis, with 3 out of 4 hospitalisations (ranging from 65.4% in Denmark to 80.7% Spain) occurring on average in children aged 0-11 months and 96% in those aged less than 0-23 months (ranging from 93.9% in Portugal to 97.7% in Norway) (Table 4).

We applied four extrapolation methods to obtain these estimates and saw no large differences across the outcomes: this was reassuring as it suggests that our results are not driven by the choice of a specific model. Consistently with previous studies, our results show an increase in RSV hospital admissions with a decrease in patient age, with infants under 1 year having the highest burden of RSV hospitalisations (especially those aged 0-2 months of age) [22]. Demont and colleagues reported a similar percentage (70%) of hospitalisations associated with RSV occurred in children <1 year [4] compared to our estimate for France (79%) and the EU-28 (75%; Table 4). Glatman-Freedman et al. (Israel), Saravanos et al. (Australia) and Arriola et al. (United States) have also found the highest age-specific hospitalisation rates in children aged 0-2 months, with reductions in the other age groups [23–25]. This confirms how RSV immunisation programmes targeting the first 6 months of life could be highly effective in reducing most of the RSV hospitalisation burden [2].

Our estimates (and specifically the hospitalisation rates) varied strongly across the different EU countries, with the Netherlands having the lowest rates in almost all age groups (Norway has the lowest rates in the age group 3-4 years) and France having the highest rates, with the highest relative difference observed in the age group 12-35 months (the estimated rate for France was 5 times higher than the Netherlands). This finding is not entirely surprising as these results reflect the Stage 1 data inputs that were entered into the Stage 2 modelling procedure, where the Netherlands had the lowest [7] and France the highest rates [15]. Differences in the outcome coding and in the study design (the French study was conducted during the winter season, and we, therefore, needed to recalculate the estimates for a whole year) may explain the higher rates reported in France. From a methodological perspective, these results highlight the importance of having Stage 1 estimates that are calculated in a harmonized manner as the Stage 2 extrapolations are sensitive to the Stage 1 inputs [26].

Whilst it is important to properly understand the real burden of disease associated with RSV in Europe and to estimate the potential impact of prevention efforts, it is not easy to compare our country-specific results with findings from the literature, considering the use of other methods to calculate these rates and the paucity of published studies. The recent, large perspective study by Wildenbeest and colleagues [27], conducted in 5 European countries, reported lower but comparable hospitalisation rates (1.8% RSV-associated hospitalisation in the first year of life in healthy term-born infants, 3.3% in children <3 months). The lower rates reported by Wildenbeest and colleagues might be related to the exclusion of pre-term infants or those at highest risk for severe illness [28] which were included in the studies used in our analysis [7, 14–15]. Sanchez-Luna and colleagues reported between 5,997 (2005) and 8,637 (2012) hospital discharges for RSV bronchiolitis (ICD-9 code 466.11 as the principal diagnosis) in Spain in children aged under 1 year during 2004-2012 [28]. Our estimated number of average admissions per year for Spain in this age group was 14,446 (95%CI 12,228-16,662) and this reflects our hospitalisation estimate not being restricted to bronchiolitis, but all respiratory hospitalisations. Moreover, as reported for England by Green et al. [29], there is an observed general increase in RSV-associated admissions over the years that may be due to changes in healthcare policies (an increase in hospital bed availability or a change in the admission threshold) and this may explain the higher number of hospitalisations estimated by our study for Spain. Our study also shows how hospitalisations due to RSV in children under 5 years represent one of the leading causes of EU infant hospitalisations (Table 5): based on our estimates, up to 1 in 10 hospitalized children under 5 years of age may be associated to RSV, and this number is larger (around 4 out of 10 children in Italy, Portugal, Denmark and Finland, 1 out of 2 in Sweden) if we only consider respiratory hospitalisations (Table 5). Accurate and reliable patient-based data on hospitalisations for multiple pathogens in children under 5 years and the related cause(s) of the hospitalisation will be fundamental in assessing whether RSV is actually the leading cause of infant hospitalisations in Europe, as recently demonstrated for the United States [30].

Our study has a number of limitations: first, our extrapolations would benefit from more countries with RSV-associated estimates to populate the statistical models (e.g., additional country estimates in southern and eastern Europe); moreover, estimates based on prospectively collected data are needed to produce more reliable results (see Supplementary material). A second limitation is that the estimates used for Stage 1 are regression-based and this holds inherent uncertainties related to country-specific collection methods of laboratory data and ICD codes for hospital admissions (ICD-10 for all countries included except the study conducted in Spain, in which ICD-9-CM was used) [31]. Without uniform reporting systems and consistent coding practices, it is hard to generalize results to other countries. Despite this, whilst differences in coding can be profound when looking at a single code, they are reduced when the modelling builds on a wider range of codes (e.g., all respiratory codes, as done by Johannesen and colleagues [7]), as clinical practices and coding guidelines are less affected. Another limitation is that our estimates are based on country-specific hospitalisation rates that were calculated for different time periods (see Table 1), thus possibly influenced by differences in RSV circulation (e.g. types) over the years. Our study found substantial variation in the hospitalisation rates across Europe, but we did not explore these differences as this would require more advanced analysis methods which would include factors such as the circulation of other respiratory viruses (e.g., influenza and SARS-CoV-2), healthcare (indicators related to access and quality of healthcare or differences in the clinical practice), climatic and environmental factors [32–33]. We also used two sets of ten indicators to produce the extrapolations (see Supplementary material): these sets were chosen based on the availability of data in all included countries (e.g., Scotland and England for the UK) and are not always specific to RSV. From a statistical perspective, this point is not likely to influence the estimates (as the indicators only aim to capture variability across countries), but it would be more elegant to develop indicator sets that are better aligned with RSV, as was done for influenza [9]. For example, the inclusion of indoor and outdoor pollution, which was reported by Nenna and colleagues as a risk factor for acute bronchiolitis in infants aged less than 3 years old [34], or the rates of premature birth, average maternal age, and delayed infant vaccinations, reported by Hardelid and co-authors as risk factors associated with increased RSV hospitalisations, could be considered [35]). Finally, our extrapolations are based on a period in which COVID-19 was not present: it would be preferable to have more recent estimates to understand the impact of the COVID-19 pandemic on RSV circulation [36] and its burden in terms of infections, hospitalisations, and deaths.

Despite these limitations, our study is, to our knowledge, the first attempt to estimate the RSV hospitalisation burden in children under the age of 5 years across Europe, and in EU countries for which no estimates have been produced so far. These estimates should help optimize public health responses (e.g., the allocation of more resources to pediatric hospitals during the winter season) and support planning for future immunisation programs [37]. Moreover, they represent a benchmark to understand to understand changes in the RSV burden after the COVID-19 pandemic and in the future following the introduction of RSV immunisation programs in Europe.

### Study group members

#### The RESCEU investigators are as follows

Harish NAIR (University of Edinburgh), Harry CAMPBELL (University of Edinburgh), Philippe Beutels (Universiteit Antwerpen), Louis Bont (University Medical Center Utrecht), Andrew Pollard (University of Oxford), Peter Openshaw (Imperial College London), Federico Martinon-Torres (Servicio Galego de Saude), Terho Heikkinen (University of Turku and Turku University Hospital), Adam Meijer (National Institute for Public Health and the Environment), Thea K. Fischer (Statens Serum Institut), Maarten van den Berge (University of Groningen), Carlo Giaquinto (PENTA Foundation), Michael Abram (AstraZeneca), Kena Swanson (Pfizer), Bishoy Rizkalla (GlaxoSmithKline), Charlotte Vernhes (Sanofi Pasteur), Scott Gallichan (Sanofi Pasteur), Jeroen Aerssens (Janssen), Veena Kumar (Novavax), Eva Molero (Team-It Research)

### Financial support

This work is part of RESCEU. RESCEU has received funding from the Innovative Medicines Initiative 2 Joint Undertaking under grant agreement No 116019. This Joint Undertaking receives support from the European Union’s Horizon 2020 research and innovation programme and EFPIA. This publication only reflects the author’s view, and the JU is not responsible for any use that may be made of the information it contains herein.

### Disclaimer

Data from the Norwegian Patient Registry have been used in this publication. The interpretation and reporting of these data are the sole responsibility of the authors, and no endorsement by the Norwegian Patient Registry is intended nor should be inferred. This work reflects only the author’s views and opinions. The EC is not responsible for any use that may be made of the information it contains.

## Data Availability

All data produced in the present study are available upon reasonable request to the authors

